# PROGNOSTIC VALUE OF COMORMIDITY FOR SEVERITY OF COVID-19: A SYSTEMATIC REVIEW AND META-ANALYSIS STUDY

**DOI:** 10.1101/2020.06.11.20128835

**Authors:** Mobina Fathi, Kimia Vakili, Fatemeh Sayehmiri, Ashraf Mohamadkhani, Mohammadreza Hajiesmaeili, Mostafa Rezaei-Tavirani, Owrang Eilami

**Affiliations:** Student research committee, Faculty of Medicine, Shahid Beheshti University of Medical Sciences, Tehran, IR Iran; Digestive Disease Research Center, Tehran University of Medical Sciences, Tehran, Iran; Anesthesiology Research Center, Loghman Hakim Hospital, Shahid Beheshti University of Medical Sciences, Tehran, IR Iran; Proteomics Research Center, Faculty of Paramedical Sciences, Shahid Beheshti University of Medical Sciences, Tehran, Iran; Associate professor of Infectious disease, Department of Family Medicine, Shiraz University of Medical Science, Fars, IR. Visitant professor of University of Sao Paulo

**Keywords:** meta-analysis, hypertension, diabetes, cardiovascular disease, comorbidity

## Abstract

**Background & Aim:** With the increase in the number of COVID-19 infections, global health is facing insufficient sources; this study aimed to provide additional data regarding the clinical characteristics of patients diagnosed with COVID-19 and in particular to analyze the factors associated with disease severity, unimprovement and mortality.

**Methods:** 82 studies were included in the present meta-analysis that all of them have been published before May 1, 2020 and were found by searching through the databases Scopus and MEDLINE. The selected papers were studied and analyzed by employing the version 14 of stata software. It should be noted that, we employed I^2^ statistics for testing and verifying heterogeneity.

**Results:** 82 papers were finally chosen for this meta-analysis, including 74855 infected patients (35673 men, 31140 women). The mean age of the patients was 56.49. The results indicate the prevalence of fever 79.84 (95% CI: 75.22-84.13), cough 59.53 (95% CI: 55.35-63.65), fatigue or myalgia 33.46 (95% CI: 28.68-38.40), dyspnea 31.48 (95% CI: 25.75-37.49) and diarrhea 10.71 (95% CI: 8.20-13.49). The prevalence of the most common comorbidities were hypertension 25.10 (95% CI: 19.91-30.64), diabetes 13.48 (95% CI: 10.61-16.62), cardiovascular diseases 8.94 (95% CI: 6.99-11.10), and chronic kidney disease 3.27 (95% CI: 2.22-4.47).

**Conclusion:** The results of this study are seriously needed to effectively monitor the health of people with comorbidities (hypertension, diabetes, cardiovascular and cerebrovascular disease, coronary heart disease, and chronic kidney disease) to prevent the development of COVID-19 infection.

**Highlights:**

- The most prevalent risk factors among patients with COVID-19 were hypertension, diabetes, cardiovascular disease, and chronic kidney disease.
- The most common symptoms among individuals who had COVID-19 infection were fever, cough, fatigue or myalgia, dyspnea, and diarrhea.
- The mean age of the patients with COVID-19 infection was 56.49.
- If the patient is an elderly male with underlying diseases, he is more likely to have severe disorders or even face to death.

## Background

On December 31, 2019, a cluster of cases of pneumonia with an unknown source was reported in Wuhan, China, related to the Huanan seafood wholesale market (1, 2). Subsequently, on January 9, 2020, the China Center for Disease Control and Prevention announced its causative factor as novel severe acute respiratory syndrome coronavirus 2 (SARS-CoV-2). This global disorder, caused by SARS-CoV-2, has been named COVID-19. Just two months later, on March 11, 2020, the Director General of the World Health Organization (WHO) announced the spread of COVID-19 as a pandemic disease (3, 4).

As of May 26, 2020, a total of 5,404,512 patients with confirmed COVID-19 infection and 343,514 dead cases have been reported by WHO globally and as of the time of this writing, its number is increasing. In addition, 216 countries, areas or territories with cases of COVID-19 infection have currently been discovered worldwide (5). As the number of COVID-19 infected patients is increasing worldwide and, due to its correlation with a huge morbidity and mortality rate, it has caused a new global phobia called Coro-phobia (6). According to the results, COVID-19 infection could be diagnosed through several symptoms such as cough, fever, fatigue, diarrhea, myalgia and dyspnea (7–9).

Huang et al. evaluated the clinical features of 41 cases who had COVID-19 infection, and their result showed that 13 (32%) of them had comorbidities comprising; hypertension, chronic obstructive pulmonary disease, diabetes, and cardiovascular disease (10). In addition, W. Guan al. reported the results of 1099 cases of COVID-19 infection and their outcome showed that 261 (23.7%) of them had underlying diseases (11). These studies and similar investigations show that the complications can be considered a risk factor for harmful outcomes in patients with COVID-19 infection (12).

Furthermore, Lai et al. (13) realized that the mortality rate was associated with the country’s healthcare resources. But in many societies, the intensive care unit (ICU) and the invasive ventilator were not enough to treat critically infected patients. Consequently, clinical operators should consider the risk factors for critical diseases of COVID-19, correctly assign medical sources, identify severe patients in the early stages of the disease and adjust an appropriate treatment plan to reduce the mortality rate and improve the effectiveness of the treatment (14). Therefore, knowing the risk factors and underlying diseases in patients with COVID-19 infection is important for healthcare professionals and especially for immunocompromised people and the elderly. Subsequently, the evaluation of the prevalence of chronic diseases such as hypertension, kidney disease, cardiovascular disease and diabetes is considered as one of the most important activities to reduce the harmful outcomes in patients with COVID-19 infection (15).

Currently, we have no specific vaccines or curative drugs approved against the new COVID-19 infection, so to prevent COVID-19 infection, a systematic review and meta-analysis has been conducted on a study of the prevalence of common comorbidities by analysis of clinical risk factors resource for COVID-19.

## Methods Study selection

We searched PubMed and Scopus databases to obtain qualified studies that were published on COVID-19 until May 1, 2020. The keywords for our research included: Kidney, COVID, Hypertension, Diabetes, ((Heart) OR Cardio-), and Comorbidities. Their references were also searched in order to collect those that are related to the title.

### Inclusion and exclusion criteria

All studies evaluating the clinical risk factors, underlying diseases and comorbidities of COVID-19 have been reviewed. For our final analysis, the studies selected should report data on the prevalence of each of the risk factors for the severity of COVID-19 and its comorbidities in patients with COVID-19 infection, including hypertension, chronic kidney disease, diabetes, cardiovascular and cerebrovascular diseases, and coronary heart disease. Only human investigations were selected. The full texts of the relevant documents were assessed for exclusion and inclusion criteria. Studies that did not have adequate data, non-human studies, duplicate publications and non-English publications (except those with useful abstract data) were excluded.

### Data extraction

All the selected papers were independently evaluated and screened by two authors (M.F. and K.V.), and the disagreement was resolved through discussion. The following data were collected for the final analysis: prevalence of hypertension, chronic kidney disease, diabetes, cardiovascular and cerebrovascular diseases, and coronary heart disease and in addition the symptoms such as fever, dyspnea, diarrhea, cough, fatigue or myalgia. Since COVID-19 is a newly developed disorder and COVID-19 related studies are still ongoing, the quality of the selected studies has not been evaluated in our document.

### Statistical analysis

Because of the effect size in this study was the proportion (prevalence of hypertension, chronic kidney disease, diabetes, cardiovascular and cerebrovascular diseases, coronary heart disease); binomial distribution was employed to calculate variance for each research. The prevalence of various researches was combined using the average weight. An inverse relationship between the variance of the study and its weight was observed. The *I*^2^ index were being applied to evaluate the heterogeneity. In the case of heterogeneous studies, random effects model were used. To analyze data, STATA software (version 14) were used. The Metaprop (meta-analysis for proportion) was used in STATA and when p was near to 0 or 1. For stabilizing the variances, we used Freeman-Tukey Double Arcsine Transformation (16). This paper was performed under the approval of ethics committee of Shahid Beheshti University of Medical Sciences (IR.SBMU.RETECH.REC.1399.084).

## Results

### Research Selection

This study written according to PRISMA checklist (17). 151 studies were initially obtained through primary search on PubMed and 49 more articles were identified through Scopus. 41 0f those 200 articles were excluded because of duplication. After screening the abstract and title of all searched papers, 47 more studies were excluded. the full text of 102 remaining studies was reviewed, 20 of them were excluded due to some reasons such as review article, case reports and short reports and insufficient data. Finally, 82 articles that have been published from February 2019 to April 2020, were selected for meta-analysis (Fig. 1, Table 1).

**Table 1.**
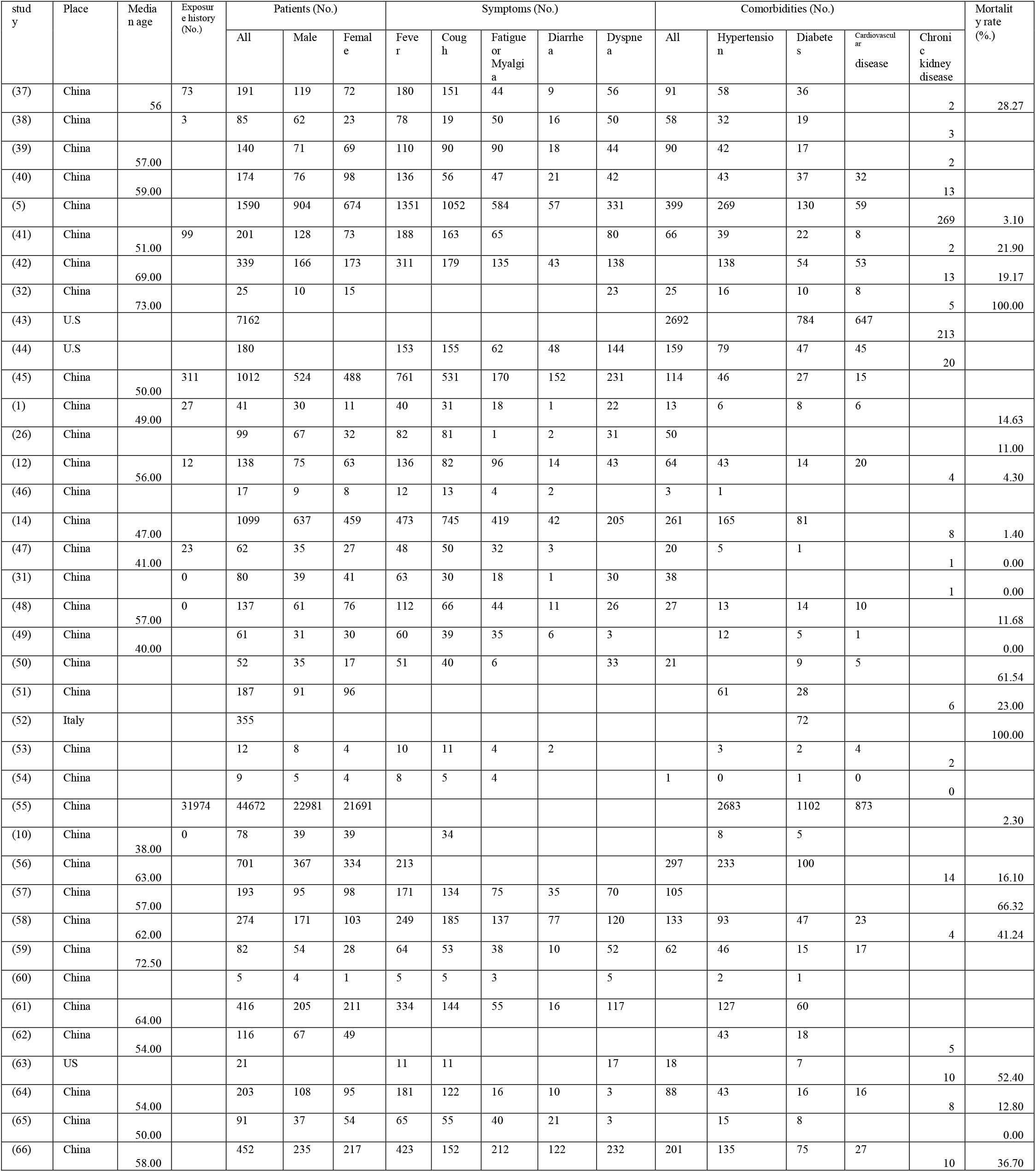
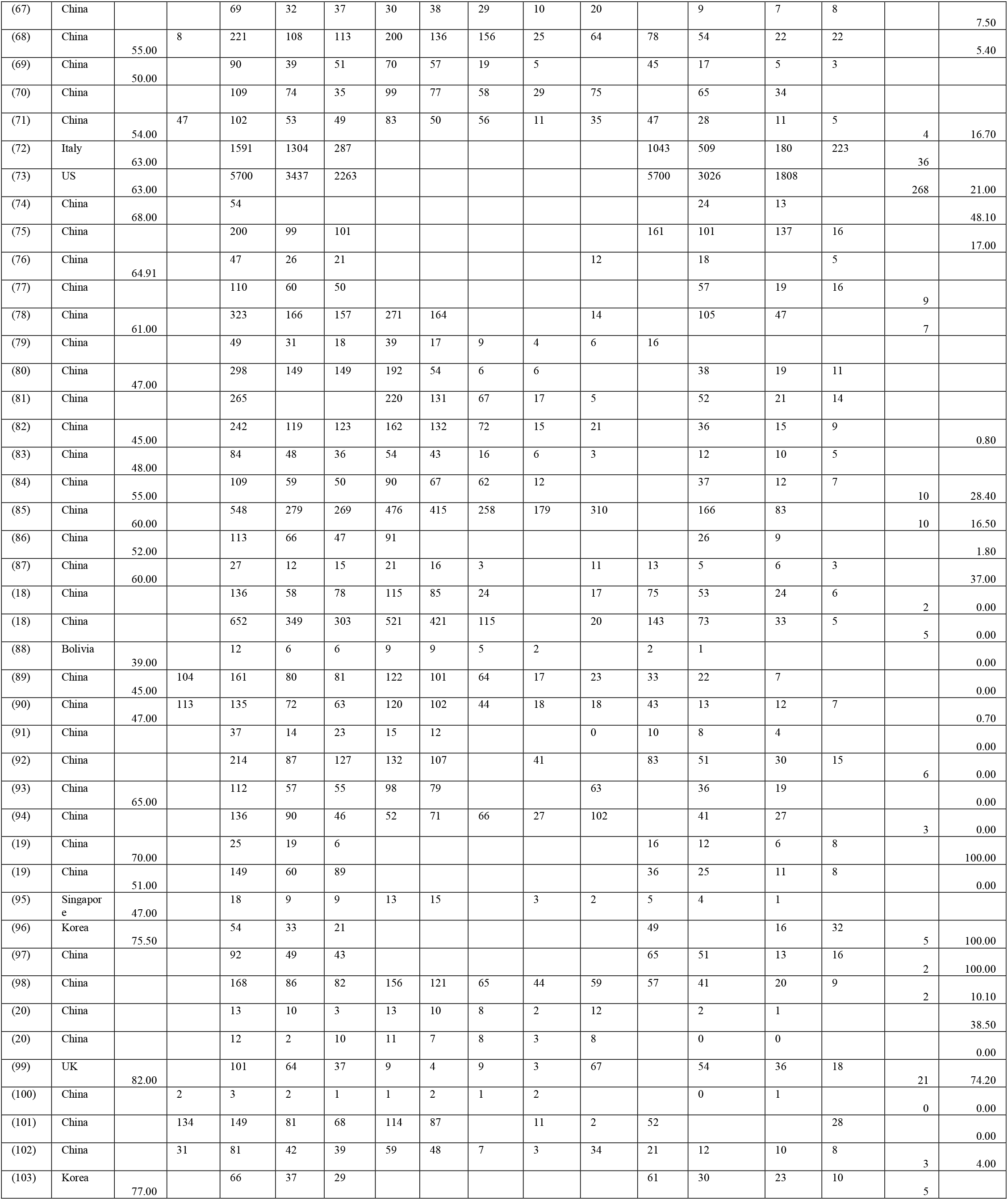
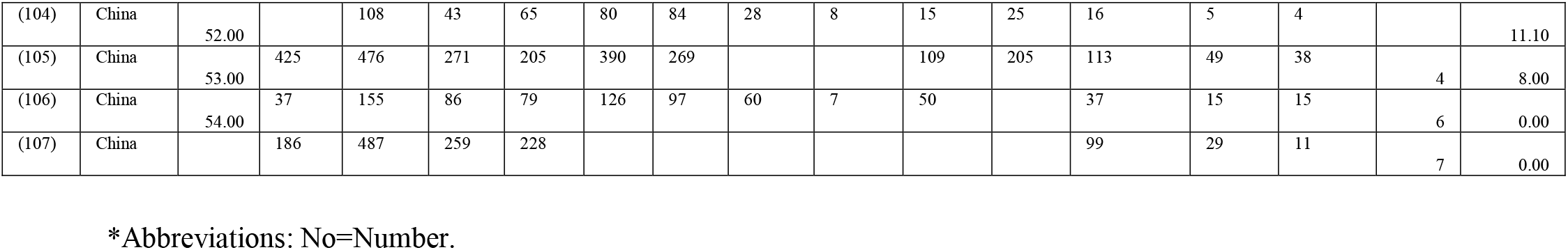
Baseline characteristics of studies included in this meta-analysis.

**Fig 1.**
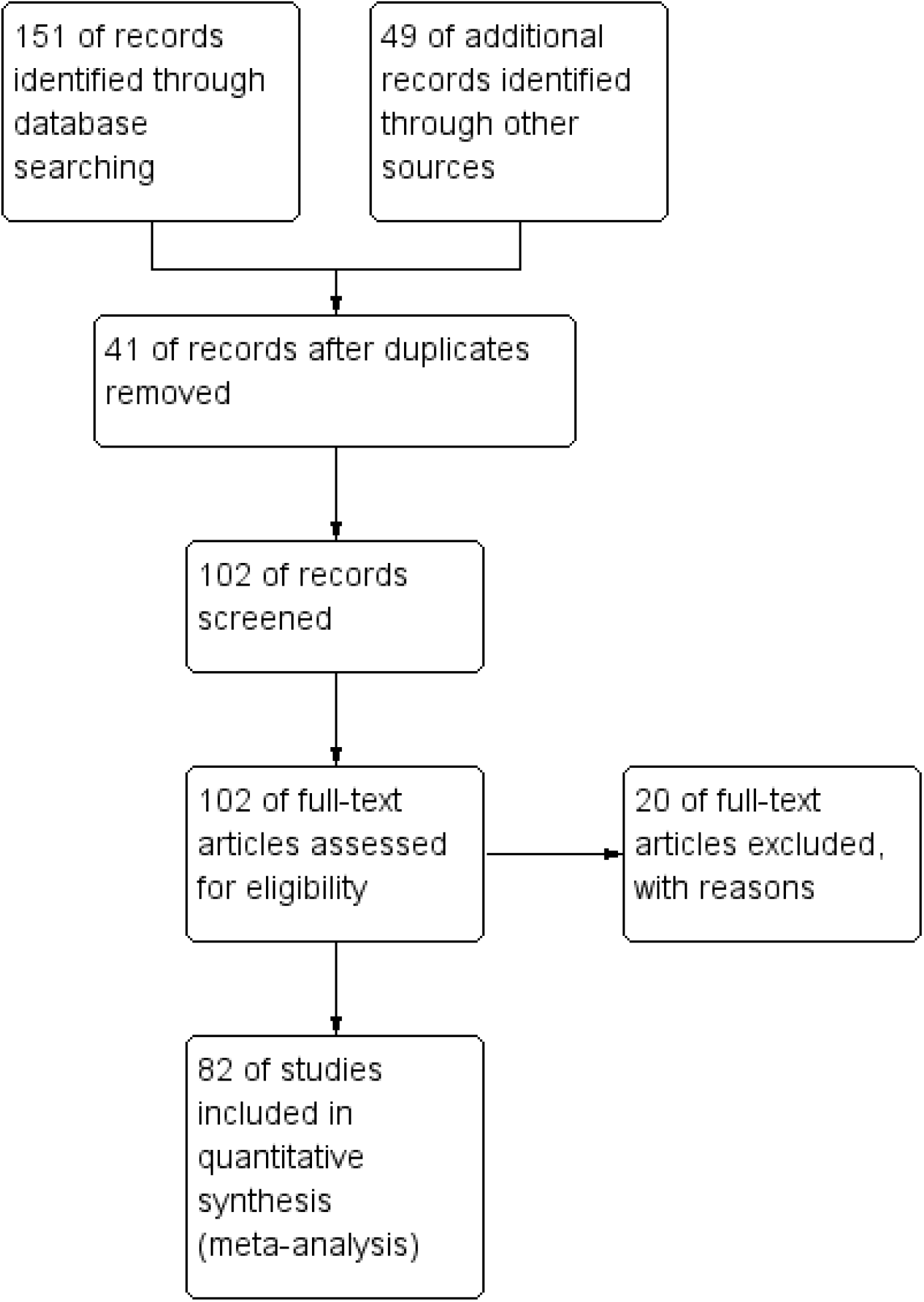
Study flow diagram.

### Demographic Characteristics

Evaluating all the data collected, the total number of hospitalized patients, including this analysis, was 74855 including 9678 patients with hypertension, 5722 patients with diabetes, 2454 patients with cardiovascular disease, 1030 patients with chronic kidney disease, 189 with coronary heart disease, 176 with cardiovascular and cerebrovascular diseases. According to results, the percentage of men was 54.26 (95% CI: 52.10-56.40; I^2^ =92.33%) and women 45.82 (95% CI: 43.83-47.98; I^2^ =92.37%) (Table 2). The mean age of the included study was 56.49. Most of our studies were conducted in China, except 4 in the United States, 2 in Italy, 2 in Korea, 1 in Bolivia and 1 in Singapore (Table2). It is important to remember that 3 (18–20) of our chosen documents divided the sample size into two groups, so we considered them as 6 studies (Table 1). We should also note that 6 of the chosen studies have reported the number of patients with both cardiovascular and cerebrovascular disease not separately. Due to papers that reported the number of individuals who had a history of exposure, the prevalence of the total exposure history was also calculated 35.56 (95% CI: 21.97-50.38; I^2^ =99.43%) (Table 2).

**Table 2.** Statistical analysis of reviewed articles

|  | Number of study | Prevalence (%) | 95% CI | I<sup>2</sup>(%) |
| --- | --- | --- | --- | --- |
| **Sex** |  |  |  |  |
| male | 79 | 54.26 | (52.10-56.40) | 92.33 |
| female | 79 | 45.82 | (43.67-47.98) | 92.37 |
| **Exposure history** | 21 | 35.56 | (21.97-50.38) | 99.43 |
| **Signs and symptoms** |  |  |  |  |
| Fever | 66 | 79.84 | (75.22-80.13) | 97.33 |
| Cough | 65 | 59.53 | (55.35-63.65) | 95.07 |
| Fatigue or Myalgia | 56 | 33.46 | (28.68-38.40) | 96.29 |
| Dyspnea | 56 | 31.48 | (25.75-37.49) | 97.80 |
| Diarrhea | 52 | 10.71 | (8.20-13.49) | 93.87 |
| **Comorbidity** | 52 | 47.01 | (35.03-59.16) | 99.69 |
| Hypertension | 75 | 25.10 | (19.91-30.64) | 99.24 |
| Cardiovascular and cerebrovascular diseases | 6 | 18.25 | (7.47-32.11) | 94.10 |
| Diabetes | 77 | 13.48 | (10.61-16.62) | 98.73 |
| Chronic kidney disease | 44 | 3.27 | (2.22-4.47) | 93.22 |
| **Heart diseases** |  |  |  |  |
| Cardiovascular disease | 51 | 8.94 | (6.99-11.10) | 97.12 |
| Coronary heart disease | 14 | 6.81 | (4.20-9.91) | 84.11 |
| **Mortality** | 55 | 8.44 | (5.86-11.38) | 98.47 |
| Mortality in China | 51 | 7.35 | (4.77-10.37) | 98.20 |
| Mortality in USA | 2 | 8.89 | (8.12-9.69) |  |
| Mortality in Bolivia | 1 | 0.00 | (0.00-26.46) |  |
| Mortality in UK | 1 | 74.26 | (64.60-82.44) |  |

### Clinical Manifestations

The results of this analysis indicates that the prevalence of the most prevalent symptoms of COVID-19 were fever 79.84 (95% CI: 75.22-84.13; I^2^ =97.33%), cough 59.53 (95% CI: 55.35-63.65; I^2^ =95.07%), fatigue or myalgia 33.46 (95% CI: 28.68-38.40; I^2^ =96.29%), dyspnea 31.48 (95% CI: 25.75-37.49; I^2^ =97.80%), and diarrhea 10.71 (95% CI: 8.20-13.49; I^2^ =93.87%) (Table 2.).

### Comorbidities

In this s systematic review, we focused on the prevalence of people with comorbidities 47.01% (95% CI: 35.03-59.16; I^2^ =99.69%) (Table 2). The prevalence of the most common comorbidities is hypertension 25.10 (95% CI: 19.91-30.64; I^2^ =99.24%) (Fig.2, Table 2), diabetes 13.48 (95% CI: 10.61-16.62; I^2^ =98.73%) (Fig.3, Table 2), cardiovascular diseases 8.94 (95% CI: 6.99-11.10; I^2^ =97.12%) (Fig.4, Table 2) and chronic kidney disease 3.27 (95% CI: 2.22-4.47; I^2^ =93.22%) (Table 2), respectively. We also assessed the prevalence of some other comorbidities found in our included articles comprising: cardiovascular and cerebrovascular diseases 18.25 (95% CI: 7.47-32.11; I^2^ =94.10%), and coronary heart disease 6.81 (95% CI: 4.20-9.91; I^2^ =84.11%) (Table 2).

**Fig 2.**
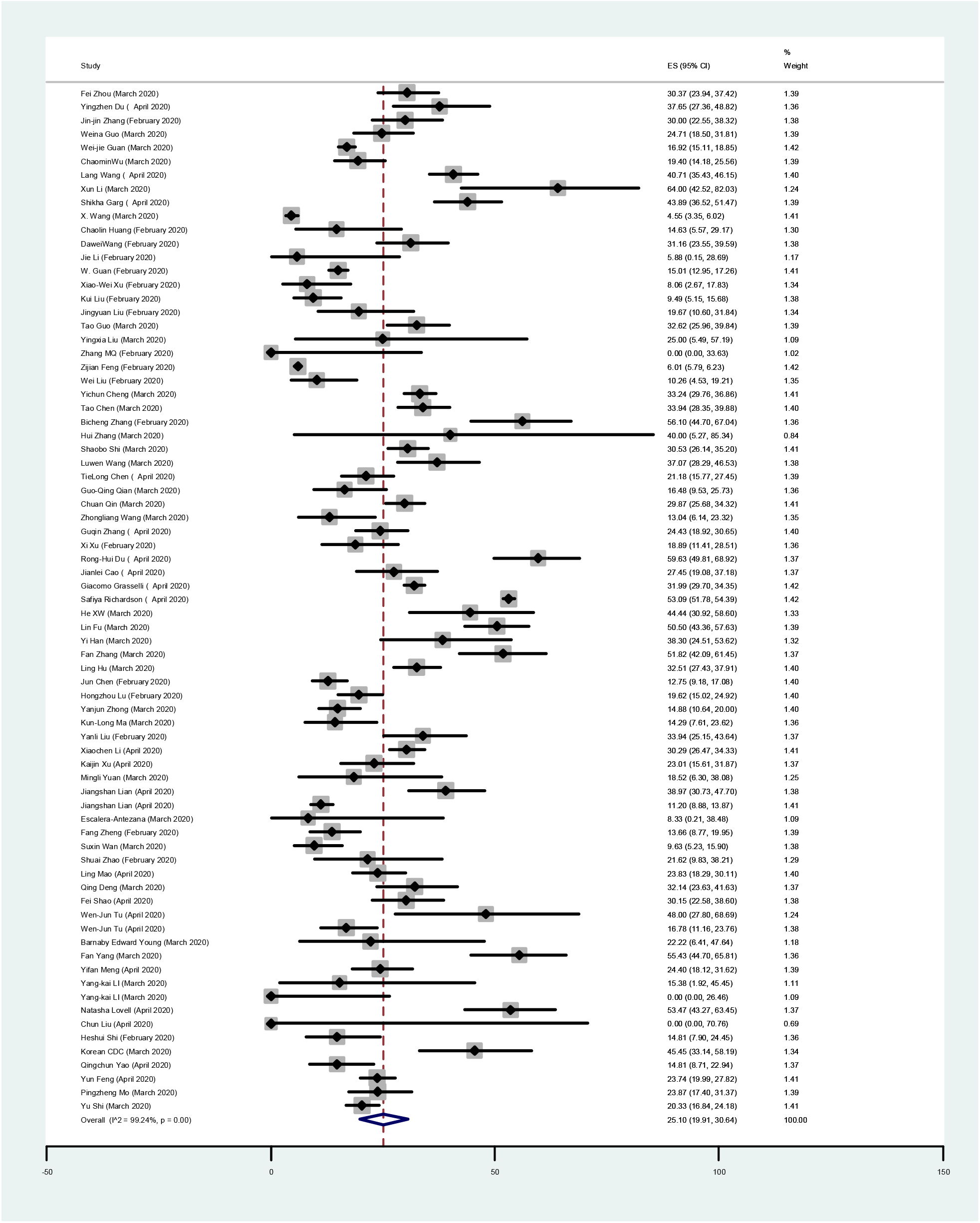
Forest Plot of the prevalence of hypertension in COVID-19 patients. Each square indicates the effect estimate of individual articles with their 95% CI Size of squares is proportional to the weight of each paper in the meta-analysis. In this plot, papers are indicated in the order of first author’s names and publication date (based on a random effects model).

**Fig 3.**
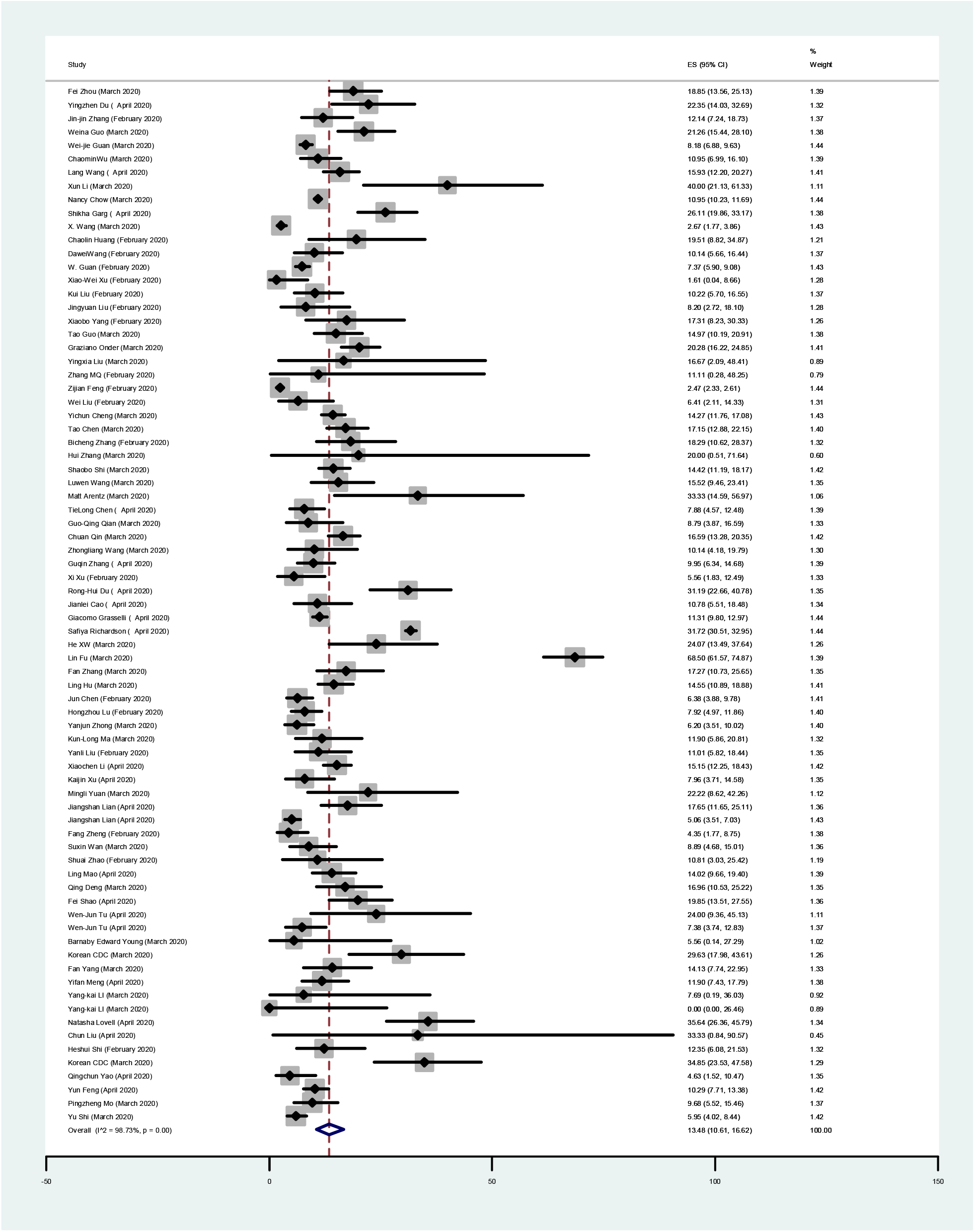
Forest Plot of the prevalence of diabetes in COVID-19 patients. Each square indicates the effect estimate of individual articles with their 95% CI Size of squares is proportional to the weight of each paper in the meta-analysis. In this plot, papers are indicated in the order of first author’s names and publication date (based on a random effects model).

**Fig 4.**
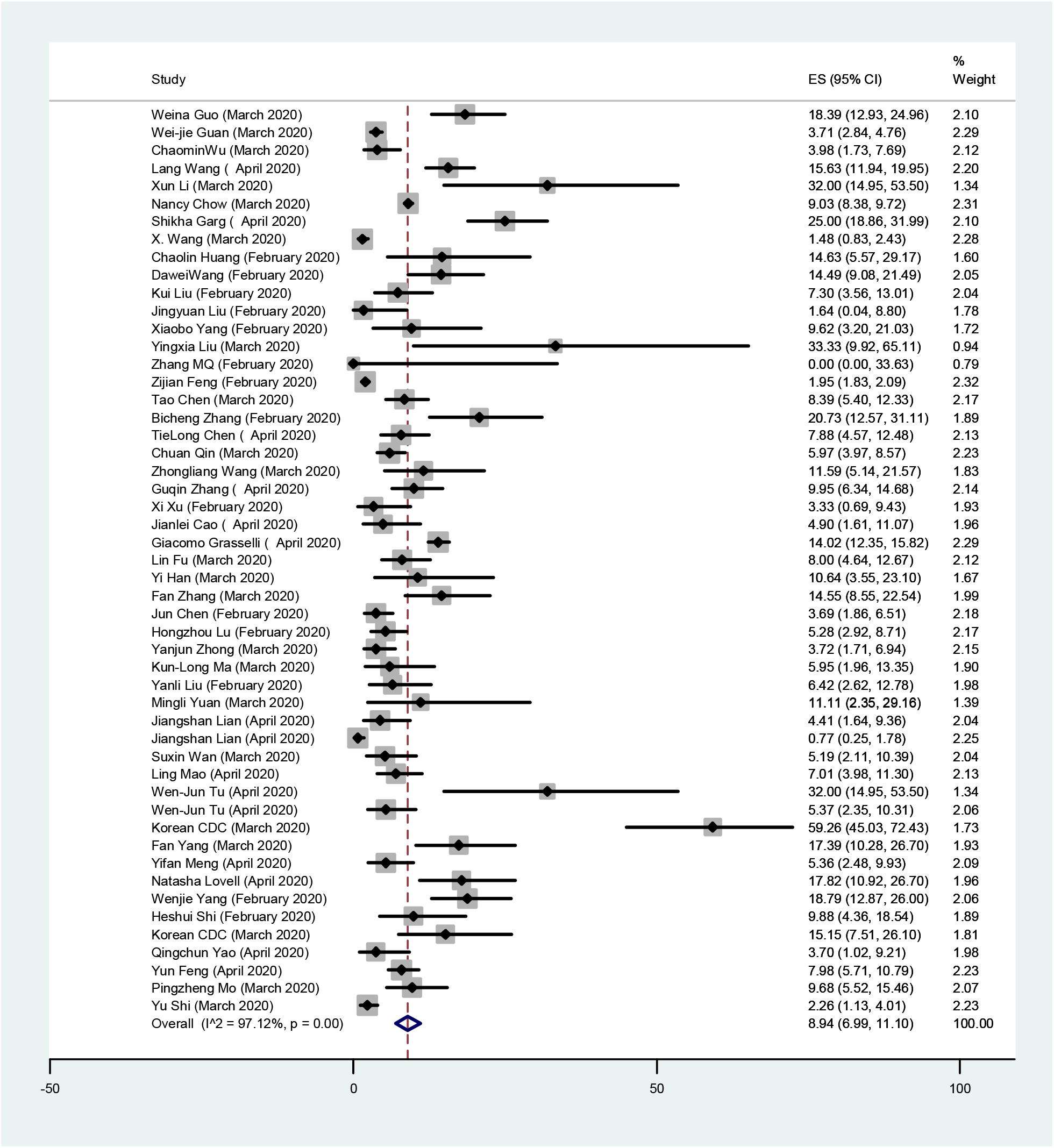
Forest Plot of the prevalence of cardiovascular disease in COVID-19 patients. Each square indicates the effect estimate of individual articles with their 95% CI Size of squares is proportional to the weight of each paper in the meta-analysis. In this plot, papers are indicated in the order of first author’s names and publication date (based on a random effects model).

### Outcomes

We calculated the percentage of the mortality rate based on all the studies reporting the case of mortality 15.01 (95% CI: 10.60-19.98; I^2^=99.21%). After removing 5 researches from our selected studies that were conducted only on deceased individuals, the overall mortality rate of the case was calculated from the remaining 55 studies 8.44 (95% CI: 5.86-11.38; I^2^=98.47%) (Table 2). After removing the studies that only assessed the deaths, a subgroup analysis was also performed to assess the prevalence of the mortality rate base on the place. As previously stated, 51 papers of our included studies were from China 7.35% (95% CI: 4.77-10.37; I^2^ =98.20%), 4 of them from USA 8.89 (95% CI: 8.12-9.69), 1 from Bolivia 0.00 (95% CI: 0.00-26.46), and 1 from UK 74.26 (95% CI: 64.60-82.44) (Table 2).

### Publication Bias

Figure 5. Shows the Beggs funnel plot of the comorbidity studies discussed in patients with COVID-19 infection. The interpretation of our Beggs funnel plot shows no sign of publication bias in included studies (p=0.240); therefore; it is understandable that reports have been published with both positive and negative outcomes (Figure 5.). Based on the results of Meta-regression analysis, the correlation between the prevalence and the sample size was compared, due to this assessment and figure, a significant relation was not observed between the prevalence and sample size (P = 0.428), also in the noted figure the circles indicate the weight of papers (Figure 6.).

**Fig 5.**
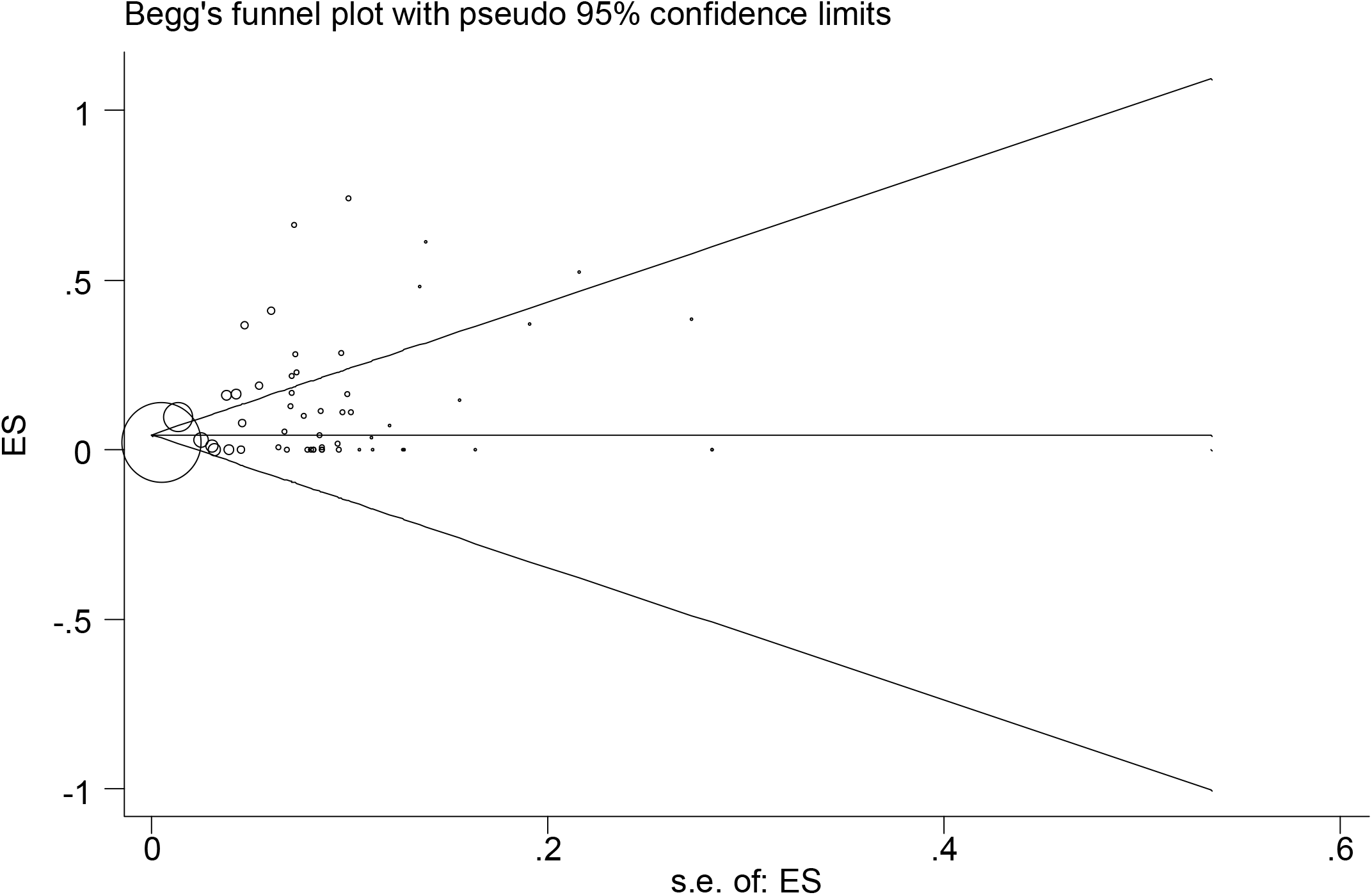
Begg’s funnel plot for publication bias

**Fig 6.**
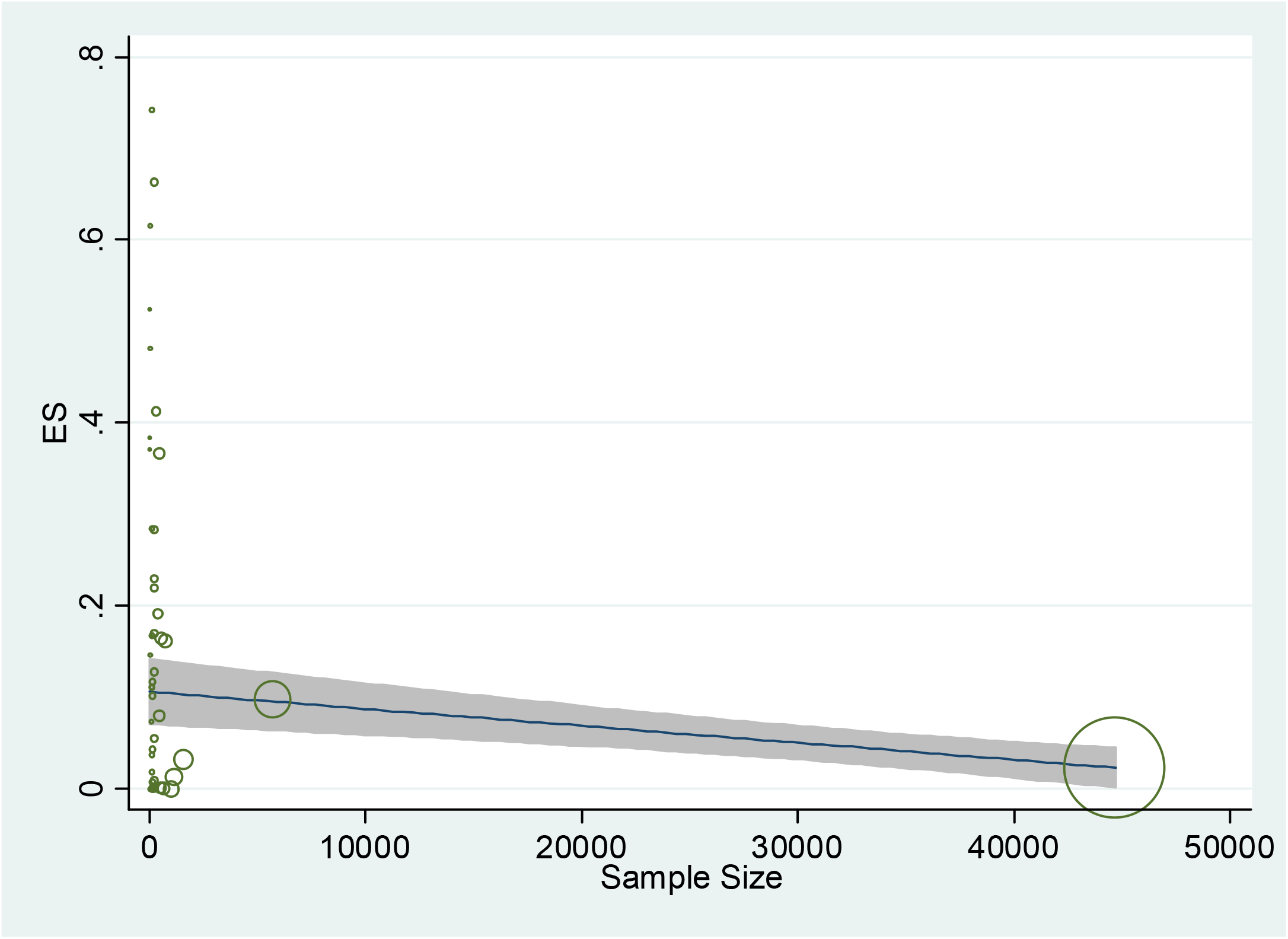
The association between prevalence of risk factors for COVID-19 and sample size, using Meta regression

## Discussion

In December 2019, a new coronavirus disease outbreak, coronavirus disease 2019 (COVID-19), spread globally (21). The city of Wuhan in China is considered to be one of the most infected areas in the world (22). Due to this epidemic contagious disease, aggressive quarantine had been suggested by the government and also played an important role in monitoring the prevalence of this virus. COVID-19 is highly contagious and insidious, it also has the ability to give rise to cluster outbreaks (23, 24). The SARS-CoV-2 virus can attack certain cells in our body, in particular alveolar epithelial cells, by a specific enzyme which is named as angiotensin-converting enzyme 2 (ACE2). ACE2 is considered to be the ACE of the isoenzyme, mainly distributed in the testes, kidneys, colon, cardiovascular system, lungs and also in other organs (25).

Up to the date of this review, the pathogenesis and origin of COVID-19 has remained unknown and no useful medical treatment has been offered for COVID-19 infection. Recommendations insist on careful and supportive care to reduce the risk of infection with this SARS-CoV-2 virus (10, 26). Unfortunately, in severely ill patients who are infected with COVID-19, the disorder develops rapidly and acute respiratory distress syndrome (ARDS) as one of the worst complications of COVID-19 outcomes can occur earlier and even lead to death. In a research by Tian et al. (27), it was stated that the percentage of mild and seriously ill patients was 73% and 18% respectively. Furthermore, Wang et al. found that the risk of exacerbation in patients with cerebrovascular disease, hypertension, cardiovascular disease, or diabetes is higher than others (1).

This meta-analysis is based on data from 82 studies with laboratory-confirmed SARS-CoV-2. The selected studies were from all over the world not a specific region. Based on previous researches on SARS-CoV and MERS-CoV, it is understood that these viruses affect males more than females (28, 29). And also because of our results, men 54.26 (95% CI: 52.10-56.40; I2 = 92.33%) are more affected with SARS-CoV-2 than women 45.82 (IC 95%: 43.83-47.98; I2 = 92.37%), moreover, this may be due to their more adaptive innate immune and robust responses (30). Another reason for our findings may be that men have a tendency to have more relationships with harmful lifestyle habits such as the underlying diseases and smoking than women (14). Moreover, as mentioned in Huang’s report (1), this may be associated with the risk factors that are related to men’s occupations with the exposure of Huanan wet market. Furthermore, due to our results, aged people (mean age=56.49) are more prone to COVID-19 and this could be related to an increase in the frequency of the common comorbidities (31). As our body’s immunity weakens with age, elderly patients are more likely to have serious ailments or even death (14).

Our study indicates the sensible evidence of risk factors in severely ill patients with COVID-19 infection, such as characteristics of the patients (gender, age), comorbidity (hypertension, chronic kidney disease, diabetes, cardiovascular and cerebrovascular diseases, and coronary heart disease) and signs and symptoms (fever, diarrhea, fatigue or myalgia and dyspnea) (32). Furthermore, we have discovered that underlying diseases such as diabetes, hypertension, cardiovascular disease, and chronic kidney disease can be considered as risk factors for the progression of this disorder. This statement conforms to the analytical results of our research. The common clinical symptoms of COVID-19 in infected patients are also fever, diarrhea, fatigue or myalgia and dyspnea and all of them have been assessed in this paper. Dyspnea or shortness of breath advises lack of oxygen and lung function. So if the patient is an elderly male with underlying diseases, he is more likely to have severe disorders or even face to death (14).

According to all of our assessed studies, we found that fever 79.84 (95% CI: 75.22-84.13; I^2^=97.33%) is the most common symptom and hypertension 25.10 (95% CI: 19.91-30.64; I^2^=99.24%) can considered as the most prevalent comorbidity. To compare our results with other recent meta-analysis studies, research conducted by Alfonso J. Rodriguez-Morales et al. (33), is evidence of our findings; they also found that fever 88.7% (95% CI 84.5–92.9) is the most prevalent symptom and hypertension 18.6 (95% CI 8.1–29.0) is most common comorbidity, however our studies were larger than they are and therefore, our findings are more reliable.

Our findings will also be approved by a study by Jing Yang et al. (15), in which fever 91.3 (95% CI: 86–97) was the most prevalent symptom and hypertension 21.1 (95% CI: 13.0–27.2) was the most common comorbidity. Besides, like a recent study by Emami et al. (34), hypertension was known as the most prevalent comorbidity 16.37 (95%CI: 10.15-23.65). According to the prevalence of hypertension and diabetes in China, diabetes mellitus and hypertension accounted for 10.9% (35) and 23.2% (36) in adults.

Regarding the studies cited and our results, individuals with cardiovascular disease, hypertension, chronic kidney disease, and diabetes should receive serious attention for treatments and vaccinations useful for SARS-CoV-2 in the future. Due to limited evidence, more appropriate studies are needed to demonstrate this correlation (15).

In summary, the results of the current review indicated that in COVID-19 patients, diabetes, hypertension, cardiovascular disease, and chronic kidney disease are the most common comorbidities and fever, diarrhea, fatigue or myalgia and dyspnea may be considered as the most prevalent symptoms among patients with COVID-19 infection. As for the fact that SARS-CoV-2 has a long incubation period and even infected people can transmit this contagious virus without indicating obvious symptoms, it is seriously suggested that sick patients who have underlying disorders, especially in epidemic areas, refrain from coming into contact with other people in society. Additionally, other strategies such as avoiding gathering in public areas and going to epidemic places should be considered as guidelines. During the current COVID-19 outbreak, the reported statistics, which are related to overall mortality, incubation time, and in particular to risk factors, imply serious decisions for future prevention and therapeutic actions (34).

## Conclusion

Due to the results of this meta-analysis, COVID-19 is notably associated with the underlying diseases. According to our results, the most frequent comorbidities among hospitalized patients with COVID-19 infection were hypertension, diabetes, cardiovascular disease and chronic kidney disease, respectively.

We also found that an old man with underlying diseases is at high risk of becoming infected with COVID-19. Therefore; given the high risk of death in patients with COVID-19 infection, special care and significant attention is needed in patients suffering from COVID-19 infection with associated conditions.

## Data Availability

available

## Acknowledgement

The authors of this study are thankful to Shahid Beheshti University of Medical Sciences,Tehran,IR Iran because of their collaboration.

No.

## Abbreviations

WHO: World Health Organization
ICU: intensive care unit
ACE2: angiotensin-converting enzyme 2
No: Number

## Declarations

## Ethics approval and consent to participate

This paper was performed under the approval of ethics committee of Shahid Beheshti University of Medical Sciences (IR.SBMU.RETECH.REC.1399.084).

## Consent for publication

The authors agree with the publication of this article.

## Availability of data and material

Not applicable.

## Authors’ contributions

Mobina Fathi: First idea, writing the manuscript, major role, editing draft Kimia Vakili: First idea, writing the manuscript, major role, editing draft Fatemeh Sayehmiri: Statistical analysis, major role, editing draft

Ashraf Mohamadkhani: scientific support

Mohammadreza Hajiesmaeili: supervision, scientific support Mostafa Rezaei-Tavirani: supervision, scientific support Owrang Eilami: editing draft

## References

1. Huang C, Wang Y, Li X, Ren L, Zhao J, Hu Y, et al. Clinical features of patients infected with 2019 novel coronavirus in Wuhan, China. The lancet. 2020;395(10223):497–506.

2. Huang Y, Lu Z, Li R, Wang B. Does comorbidity increase the risk of patients with COVID-19: evidence from metaanalysis. Aging. 2020;12(7):6049–57.

3. Organization WH. WHO announces COVID-19 outbreak a pandemic. WHO, Geneva, Switzerland. 2020.

4. Kreutz R, Algharably EAE-H, Azizi M, Dobrowolski P, Guzik T, Januszewicz A, et al. Hypertension, the renin– angiotensin system, and the risk of lower respiratory tract infections and lung injury: implications for COVID-19 European Society of Hypertension COVID-19 Task Force Review of Evidence. Cardiovascular Research. 2020.

5. Guan W-j, Liang W-h, Zhao Y, Liang H-r, Chen Z-s, Li Y-m, et al. Comorbidity and its impact on 1590 patients with Covid-19 in China: A Nationwide Analysis. European Respiratory Journal. 2020.

6. Kelvin DJ, Rubino S. Fear of the novel coronavirus. The Journal of Infection in Developing Countries. 2020;14(01):1–2.

7. Carlos WG, Dela Cruz CS, Cao B, Pasnick S, Jamil S. Novel wuhan (2019-nCoV) coronavirus. American journal of respiratory and critical care medicine. 2020;201(4):P7–P8.

8. Paules CI, Marston HD, Fauci AS. Coronavirus infections—more than just the common cold. Jama. 2020;323(8):707–8.

9. Livingston E, Bucher K, Rekito A. Coronavirus Disease 2019 and Influenza 2019-2020. Jama. 2020;323(12):1122-

10. Liu W, Tao Z-W, Wang L, Yuan M-L, Liu K, Zhou L, et al. Analysis of factors associated with disease outcomes in hospitalized patients with 2019 novel coronavirus disease. Chinese medical journal. 2020.

11. Guan W-j, Ni Z-y, Hu Y, Liang W-h, Ou C-q, He J-x, et al. Clinical characteristics of coronavirus disease 2019 in China. New England journal of medicine. 2020.

12. Wang D, Hu B, Hu C, Zhu F, Liu X, Zhang J, et al. Clinical characteristics of 138 hospitalized patients with 2019 novel coronavirus–infected pneumonia in Wuhan, China. Jama. 2020;323(11):1061–9.

13. Lai C-C, Wang C-Y, Wang Y-H, Hsueh S-C, Ko W-C, Hsueh P-R. Global epidemiology of coronavirus disease 2019: disease incidence, daily cumulative index, mortality, and their association with country healthcare resources and economic status. International Journal of Antimicrobial Agents. 2020:105946.

14. Guan W-j, Ni Z-y, Hu Y, Liang W-h, Ou C-q, He J-x, et al. Clinical characteristics of coronavirus disease 2019 in China. New England journal of medicine. 2020;382(18):1708–20.

15. Yang J, Zheng Y, Gou X, Pu K, Chen Z, Guo Q, et al. Prevalence of comorbidities in the novel Wuhan coronavirus (COVID-19) infection: a systematic review and meta-analysis. International Journal of Infectious Diseases. 2020.

16. Freeman MF, Tukey JW. Transformations related to the angular and the square root. The Annals of Mathematical Statistics. 1950:607–11.

17. Liberati A, Altman DG, Tetzlaff J, Mulrow C, Gøtzsche PC, Ioannidis JP, et al. The PRISMA statement for reporting systematic reviews and meta-analyses of studies that evaluate health care interventions: explanation and elaboration. Annals of internal medicine. 2009;151(4):W-65-W-94.

18. Lian J, Jin X, Hao S, Cai H, Zhang S, Zheng L, et al. Analysis of Epidemiological and Clinical features in older patients with Corona Virus Disease 2019 (COVID-19) out of Wuhan. Clinical Infectious Diseases. 2020.

19. Tu W-J, Cao J, Yu L, Hu X, Liu Q. Clinicolaboratory study of 25 fatal cases of COVID-19 in Wuhan. Intensive care medicine. 2020:1–4.

20. Li Y-K, Peng S, Li L-Q, Wang Q, Ping W, Zhang N, et al. Clinical and transmission characteristics of Covid-19—a retrospective study of 25 cases from a single thoracic surgery department. Current Medical Science. 2020:1–6.

21. Ghebreyesus TA. WHO Director-General’s opening remarks at the media briefing on COVID-19-11 March 2020. World Health Organization https://www.who.int/dg/speeches/detail/who-director-general-s-opening-remarks-atthe-media-briefing-on-covid-19---11-march-2020. 2020.

22. Li J, Xu G. Lessons from the Experience in Wuhan to Reduce Risk of COVID-19 Infection in Patients Undergoing Long-Term Hemodialysis. Clinical Journal of the American Society of Nephrology. 2020.

23. Chan JF-W, Yuan S, Kok K-H, To KK-W, Chu H, Yang J, et al. A familial cluster of pneumonia associated with the 2019 novel coronavirus indicating person-to-person transmission: a study of a family cluster. The Lancet. 2020;395(10223):514–23.

24. Guan W, Ni Z, Hu Y, Liang W, Ou C, He J, et al. China Medical Treatment Expert Group for Covid-19. Clinical characteristics of coronavirus disease. 2019.

25. Tipnis S. Hooper NM, Hyde R, Karran E, Christie G, Turner AJ. A human homolog of angiotensin-converting enzyme Cloning and functional expression as a captopril-insensitive carboxypeptidase J Biol Chem. 2000;275:33238–43.

26. Chen N, Zhou M, Dong X, Qu J, Gong F, Han Y, et al. Epidemiological and clinical characteristics of 99 cases of 2019 novel coronavirus pneumonia in Wuhan, China: a descriptive study. The Lancet. 2020;395(10223):507–13.

27. Tian S, Hu N, Lou J, Chen K, Kang X, Xiang Z, et al. Characteristics of COVID-19 infection in Beijing. Journal of Infection. 2020.

28. Channappanavar R, Fett C, Mack M, Ten Eyck PP, Meyerholz DK, Perlman S. Sex-based differences in susceptibility to severe acute respiratory syndrome coronavirus infection. The Journal of Immunology. 2017;198(10):4046–53.

29. Badawi A, Ryoo SG. Prevalence of comorbidities in the Middle East respiratory syndrome coronavirus (MERS-CoV): a systematic review and meta-analysis. International Journal of Infectious Diseases. 2016;49:129–33.

30. Jaillon S, Berthenet K, Garlanda C. Sexual dimorphism in innate immunity. Clinical reviews in allergy & immunology. 2017:1–14.

31. Wu J, Liu J, Zhao X, Liu C, Wang W, Wang D, et al. Clinical Characteristics of Imported Cases of Coronavirus Disease 2019 (COVID-19) in Jiangsu Province: A Multicenter Descriptive Study. Clinical Infectious Diseases. 2020.

32. Li X, Wang L, Yan S, Yang F, Xiang L, Zhu J, et al. Clinical characteristics of 25 death cases with COVID-19: a retrospective review of medical records in a single medical center, Wuhan, China. International Journal of Infectious Diseases. 2020.

33. Rodriguez-Morales AJ, Cardona-Ospina JA, Gutiérrez-Ocampo E, Villamizar-Peña R, Holguin-Rivera Y, Escalera-Antezana JP, et al. Clinical, laboratory and imaging features of COVID-19: A systematic review and meta-analysis. Travel medicine and infectious disease. 2020:101623.

34. Emami A, Javanmardi F, Pirbonyeh N, Akbari A. Prevalence of underlying diseases in hospitalized patients with COVID-19: a systematic review and meta-analysis. Archives of Academic Emergency Medicine. 2020;8(1).

35. Liu M, Liu S-W, Wang L-J, Bai Y-M, Zeng X-Y, Guo H-B, et al. Burden of diabetes, hyperglycaemia in China from to 2016: findings from the 1990 to 2016, global burden of disease study. Diabetes & metabolism. 2019;45(3):286–93.

36. Hu S, Gao R, Liu L, Zhu M, Wang W, Wang Y, et al. Summary of the 2018 report on cardiovascular diseases in China. Chin Circulation J. 2019;34:209.

37. Zhou F, Yu T, Du R, Fan G, Liu Y, Liu Z, et al. Clinical course and risk factors for mortality of adult inpatients with COVID-19 in Wuhan, China: a retrospective cohort study. The lancet. 2020.

38. Du Y, Tu L, Zhu P, Mu M, Wang R, Yang P, et al. Clinical features of 85 fatal cases of COVID-19 from Wuhan: a retrospective observational study. American journal of respiratory and critical care medicine. 2020(ja).

39. Zhang J-j, Dong X, Cao Y-y, Yuan Y-d, Yang Y-b, Yan Y-q, et al. Clinical characteristics of 140 patients infected with SARS-CoV-2 in Wuhan, China. Allergy. 2020.

40. Guo W, Li M, Dong Y, Zhou H, Zhang Z, Tian C, et al. Diabetes is a risk factor for the progression and prognosis of COVID-19. Diabetes/metabolism research and reviews. 2020.

41. Wu C, Chen X, Cai Y, Zhou X, Xu S, Huang H, et al. Risk factors associated with acute respiratory distress syndrome and death in patients with coronavirus disease 2019 pneumonia in Wuhan, China. JAMA internal medicine. 2020.

42. Wang L, He W, Yu X, Hu D, Bao M, Liu H, et al. Coronavirus Disease 2019 in elderly patients: characteristics and prognostic factors based on 4-week follow-up. Journal of Infection. 2020.

43. Chow N, Fleming-Dutra K, Gierke R, Hall A, Hughes M, Pilishvili T, et al. Preliminary estimates of the prevalence of selected underlying health conditions among patients with coronavirus disease 2019—United States, February 12 – March 28, 2020. 2020.

44. Garg S. Hospitalization rates and characteristics of patients hospitalized with laboratory-confirmed coronavirus disease 2019—COVID-NET, 14 States, March 1–30, 2020. MMWR Morbidity and Mortality Weekly Report. 2020;69.

45. Wang X, Fang J, Zhu Y, Chen L, Ding F, Zhou R, et al. Clinical characteristics of non-critically ill patients with novel coronavirus infection (COVID-19) in a Fangcang Hospital. Clinical Microbiology and Infection. 2020.

46. Li J, Li S, Cai Y, Liu Q, Li X, Zeng Z, et al. Epidemiological and Clinical Characteristics of 17 Hospitalized Patients with 2019 Novel Coronavirus Infections Outside Wuhan, China. medRxiv. 2020.

47. Xu X-W, Wu X-X, Jiang X-G, Xu K-J, Ying L-J, Ma C-L, et al. Clinical findings in a group of patients infected with the 2019 novel coronavirus (SARS-Cov-2) outside of Wuhan, China: retrospective case series. bmj. 2020;368.

48. Liu K, Fang Y-Y, Deng Y, Liu W, Wang M-F, Ma J-P, et al. Clinical characteristics of novel coronavirus cases in tertiary hospitals in Hubei Province. Chinese medical journal. 2020.

49. Liu J, Liu Y, Xiang P, Pu L, Xiong H, Li C, et al. Neutrophil-to-lymphocyte ratio predicts severe illness patients with 2019 novel coronavirus in the early stage. MedRxiv. 2020.

50. Yang X, Yu Y, Xu J, Shu H, Liu H, Wu Y, et al. Clinical course and outcomes of critically ill patients with SARS-CoV-2 pneumonia in Wuhan, China: a single-centered, retrospective, observational study. The Lancet Respiratory Medicine. 2020.

51. Guo T, Fan Y, Chen M, Wu X, Zhang L, He T, et al. Cardiovascular implications of fatal outcomes of patients with coronavirus disease 2019 (COVID-19). JAMA cardiology. 2020.

52. Onder G, Rezza G, Brusaferro S. Case-fatality rate and characteristics of patients dying in relation to COVID-19 in Italy. Jama. 2020.

53. Liu Y, Yang Y, Zhang C, Huang F, Wang F, Yuan J, et al. Clinical and biochemical indexes from 2019-nCoV infected patients linked to viral loads and lung injury. Science China Life Sciences. 2020;63(3):364–74.

54. Zhang MQ, Wang XH, Chen YL, Zhao KL, Cai YQ, An CL, et al. [Clinical features of 2019 novel coronavirus pneumonia in the early stage from a fever clinic in Beijing]. Zhonghua jie he he hu xi za zhi = Zhonghua jiehe he huxi zazhi = Chinese journal of tuberculosis and respiratory diseases. 2020;43(0):E013.

55. Surveillances V. The epidemiological characteristics of an outbreak of 2019 novel coronavirus diseases (COVID-19)—China, 2020. China CDC Weekly. 2020;2(8):113–22.

56. Cheng Y, Luo R, Wang K, Zhang M, Wang Z, Dong L, et al. Kidney disease is associated with in-hospital death of patients with COVID-19. Kidney International. 2020.

57. Li Z, Wu M, Yao J, Guo J, Liao X, Song S, et al. Caution on kidney dysfunctions of COVID-19 patients. 2020.

58. Chen T, Wu D, Chen H, Yan W, Yang D, Chen G, et al. Clinical characteristics of 113 deceased patients with coronavirus disease 2019: retrospective study. Bmj. 2020;368.

59. Zhang B, Zhou X, Qiu Y, Feng F, Feng J, Jia Y, et al. Clinical characteristics of 82 death cases with COVID-19. medRxiv. 2020.

60. Zhang H, Chen Y, Yuan Q, Xia Q-X, Zeng X-P, Peng J-T, et al. Identification of kidney transplant recipients with coronavirus disease 2019. European Urology. 2020.

61. Shi S, Qin M, Shen B, Cai Y, Liu T, Yang F, et al. Association of cardiac injury with mortality in hospitalized patients with COVID-19 in Wuhan, China. JAMA cardiology. 2020.

62. Wang L, Li X, Chen H, Yan S, Li D, Li Y, et al. Coronavirus disease 19 infection does not result in acute kidney injury: an analysis of 116 hospitalized patients from Wuhan, China. American journal of nephrology. 2020;51(5):343–8.

63. Arentz M, Yim E, Klaff L, Lokhandwala S, Riedo FX, Chong M, et al. Characteristics and outcomes of 21 critically ill patients with COVID-19 in Washington State. Jama. 2020.

64. Chen T, Dai Z, Mo P, Li X, Ma Z, Song S, et al. Clinical characteristics and outcomes of older patients with coronavirus disease 2019 (COVID-19) in Wuhan, China (2019): a single-centered, retrospective study. The Journals of Gerontology: Series A. 2020.

65. Qian G-Q, Yang N-B, Ding F, Ma AHY, Wang Z-Y, Shen Y-F, et al. Epidemiologic and Clinical Characteristics of 91 Hospitalized Patients with COVID-19 in Zhejiang, China: A retrospective, multi-centre case series. QJM: An International Journal of Medicine. 2020.

66. Qin C, Zhou L, Hu Z, Zhang S, Yang S, Tao Y, et al. Dysregulation of immune response in patients with COVID-19 in Wuhan, China. Clinical Infectious Diseases. 2020.

67. Wang Z, Yang B, Li Q, Wen L, Zhang R. Clinical Features of 69 Cases with Coronavirus Disease 2019 in Wuhan, China. Clinical infectious diseases : an official publication of the Infectious Diseases Society of America. 2020.

68. Zhang G, Hu C, Luo L, Fang F, Chen Y, Li J, et al. Clinical features and short-term outcomes of 221 patients with COVID-19 in Wuhan, China. Journal of Clinical Virology. 2020:104364.

69. Xu X, Yu C, Qu J, Zhang L, Jiang S, Huang D, et al. Imaging and clinical features of patients with 2019 novel coronavirus SARS-CoV-2. European journal of nuclear medicine and molecular imaging. 2020:1–6.

70. Du R-H, Liu L-M, Yin W, Wang W, Guan L-L, Yuan M-L, et al. Hospitalization and Critical Care of 109 Decedents with COVID-19 Pneumonia in Wuhan, China. Annals of the American Thoracic Society. 2020(ja).

71. Cao J, Tu W-J, Cheng W, Yu L, Liu Y-K, Hu X, et al. Clinical Features and Short-term Outcomes of 102 Patients with Corona Virus Disease 2019 in Wuhan, China. Clinical Infectious Diseases. 2020.

72. Grasselli G, Zangrillo A, Zanella A, Antonelli M, Cabrini L, Castelli A, et al. Baseline characteristics and outcomes of 1591 patients infected with SARS-CoV-2 admitted to ICUs of the Lombardy Region, Italy. Jama. 2020.

73. Richardson S, Hirsch JS, Narasimhan M, Crawford JM, McGinn T, Davidson KW, et al. Presenting Characteristics, Comorbidities, and Outcomes Among 5700 Patients Hospitalized With COVID-19 in the New York City Area. JAMA. 2020.

74. He XW, Lai JS, Cheng J, Wang MW, Liu YJ, Xiao ZC, et al. [Impact of complicated myocardial injury on the clinical outcome of severe or critically ill COVID-19 patients]. Zhonghua xin xue guan bing za zhi. 2020;48(0):E011.

75. Fu L, Fei J, Xiang H-X, Xiang Y, Tan Z-X, Li M-D, et al. Influence factors of death risk among COVID-19 patients in Wuhan, China: a hospital-based case-cohort study. medRxiv. 2020:2020.03.13.20035329.

76. Han Y, Zhang H, Mu S, Wei W, Jin C, Xue Y, et al. Lactate dehydrogenase, a Risk Factor of Severe COVID-19 Patients. medRxiv. 2020.

77. Zhang F, Yang D, Li J, Gao P, Chen T, Cheng Z, et al. Myocardial injury is associated with in-hospital mortality of confirmed or suspected COVID-19 in Wuhan, China: A single center retrospective cohort study. MedRxiv. 2020.

78. Hu L, Chen S, Fu Y, Gao Z, Long H, Ren H-w, et al. Risk factors associated with clinical outcomes in 323 COVID-19 patients in Wuhan, China. Medrxiv. 2020.

79. Ji D, Zhang D, Chen Z, Xu Z, Zhao P, Zhang M, et al. Clinical Characteristics Predicting Progression of COVID-19. 2020.

80. Cai Q, Huang D, Ou P, Yu H, Zhu Z, Xia Z, et al. 2019-nCoV Pneumonia in a Normal Work Infectious Diseases Hospital Besides Hubei Province, China. 2020.

81. Lu H, Ai J, Shen Y, Li Y, Li T, Zhou X, et al. A descriptive study of the impact of diseases control and prevention on the epidemics dynamics and clinical features of SARS-CoV-2 outbreak in Shanghai, lessons learned for metropolis epidemics prevention. medRxiv. 2020.

82. Wang G, Wu C, Zhang Q, Wu F, Yu B, Lv J, et al. Epidemiological and Clinical Features of Corona Virus Disease 2019 (COVID-19) in Changsha, China. China (3/1/2020). 2020.

83. Ma K-L, Liu Z-H, Cao C-f, Liu M-K, Liao J, Zou J-B, et al. COVID-19 myocarditis and severity factors: an adult cohort study. medRxiv. 2020.

84. Liu Y, Sun W, Li J, Chen L, Wang Y, Zhang L, et al. Clinical features and progression of acute respiratory distress syndrome in coronavirus disease 2019. MedRxiv. 2020.

85. Li X, Xu S, Yu M, Wang K, Tao Y, Zhou Y, et al. Risk factors for severity and mortality in adult COVID-19 inpatients in Wuhan. Journal of Allergy and Clinical Immunology. 2020.

86. Xu K, Chen Y, Yuan J, Yi P, Ding C, Wu W, et al. Factors associated with prolonged viral RNA shedding in patients with COVID-19. Clinical Infectious Diseases. 2020.

87. Yuan M, Yin W, Tao Z, Tan W, Hu Y. Association of radiologic findings with mortality of patients infected with 2019 novel coronavirus in Wuhan, China. PLoS One. 2020;15(3):e0230548.

88. Escalera-Antezana JP, Lizon-Ferrufino NF, Maldonado-Alanoca A, Alarcón-De-la-Vega G, Alvarado-Arnez LE, Balderrama-Saavedra MA, et al. Clinical features of cases and a cluster of Coronavirus Disease 2019 (COVID-19) in Bolivia imported from Italy and Spain. Travel Medicine and Infectious Disease. 2020:101653.

89. Zheng F, Tang W, Li H, Huang Y, Xie Y, Zhou Z. Clinical characteristics of 161 cases of corona virus disease 2019 (COVID-19) in Changsha. Eur Rev Med Pharmacol Sci. 2020;24(6):3404–10.

90. Wan S, Xiang Y, Fang W, Zheng Y, Li B, Hu Y, et al. Clinical Features and Treatment of COVID-19 Patients in Northeast Chongqing. Journal of medical virology. 2020.

91. Zhao S, Ling K, Yan H, Zhong L, Peng X, Yao S, et al. Anesthetic management of patients with suspected or confirmed 2019 novel coronavirus infection during emergency procedures. J Cardiothorac Vasc Anesth. 2020;28:28.

92. Mao L, Jin H, Wang M, Hu Y, Chen S, He Q, et al. Neurologic manifestations of hospitalized patients with coronavirus disease 2019 in Wuhan, China. JAMA neurology. 2020.

93. Deng Q, Hu B, Zhang Y, Wang H, Zhou X, Hu W, et al. Suspected myocardial injury in patients with COVID-19: Evidence from front-line clinical observation in Wuhan, China. International Journal of Cardiology. 2020.

94. Shao F, Xu S, Ma X, Xu Z, Lyu J, Ng M, et al. In-hospital cardiac arrest outcomes among patients with COVID-19 pneumonia in Wuhan, China. Resuscitation. 2020.

95. Young BE, Ong SWX, Kalimuddin S, Low JG, Tan SY, Loh J, et al. Epidemiologic features and clinical course of patients infected with SARS-CoV-2 in Singapore. Jama. 2020;323(15):1488–94.

96. Park S, Lee M, Kim S, Kwak Y, Kwon K, Park J, et al. Analysis on 54 Mortality Cases of Coronavirus Disease 2019 in the Republic of Korea from January 19 to March 10, 2020. J Korean Med Sci. 2020;35(12):e132.

97. Yang F, Shi S, Zhu J, Shi J, Dai K, Chen X. Analysis of 92 deceased patients with COVID-19. Journal of Medical Virology. 2020.

98. Meng Y, Wu P, Lu W, Liu K, Ma K, Huang L, et al. Sex-specific clinical characteristics and prognosis of coronavirus disease-19 infection in Wuhan, China: A retrospective study of 168 severe patients. PLoS pathogens. 2020;16(4):e1008520.

99. Lovell N, Maddocks M, Etkind SN, Taylor K, Carey I, Vora V, et al. Characteristics, symptom management and outcomes of 101 patients with COVID-19 referred for hospital palliative care. Journal of Pain and Symptom Management. 2020.

100. Liu C, Wu C, Zheng X, Zeng F, Liu J, Wang P, et al. Clinical features and multidisciplinary treatment outcome of COVID-19 pneumonia: A report of three cases. Journal of the Formosan Medical Association. 2020.

101. Yang W, Cao Q, Qin L, Wang X, Cheng Z, Pan A, et al. Clinical characteristics and imaging manifestations of the 2019 novel coronavirus disease (COVID-19): A multi-center study in Wenzhou city, Zhejiang, China. Journal of Infection. 2020.

102. Shi H, Han X, Jiang N, Cao Y, Alwalid O, Gu J, et al. Radiological findings from 81 patients with COVID-19 pneumonia in Wuhan, China: a descriptive study. The Lancet Infectious Diseases. 2020.

103. Choe YJ. Coronavirus disease-19: The First 7,755 Cases in the Republic of Korea. medRxiv. 2020.

104. Yao Q, Wang P, Wang X, Qie G, Meng M, Tong X, et al. Retrospective study of risk factors for severe SARS-Cov-2 infections in hospitalized adult patients. Polish archives of internal medicine. 2020.

105. Feng Y, Ling Y, Bai T, Xie Y, Huang J, Li J, et al. COVID-19 with Different Severity: A Multi-center Study of Clinical Features. American Journal of Respiratory and Critical Care Medicine. 2020(ja).

106. Mo P, Xing Y, Xiao Y, Deng L, Zhao Q, Wang H, et al. Clinical characteristics of refractory COVID-19 pneumonia in Wuhan, China. Clinical Infectious Diseases. 2020.

107. Shi Y, Yu X, Zhao H, Wang H, Zhao R, Sheng J. Host susceptibility to severe COVID-19 and establishment of a host risk score: findings of 487 cases outside Wuhan. Critical Care. 2020;24(1):1–4.

